# Bounce backs amid continued losses: Life expectancy changes since COVID-19

**DOI:** 10.1101/2022.02.23.22271380

**Authors:** Jonas Schöley, José Manuel Aburto, Ilya Kashnitsky, Maxi S. Kniffka, Luyin Zhang, Hannaliis Jaadla, Jennifer B. Dowd, Ridhi Kashyap

## Abstract

The COVID-19 pandemic triggered an unprecedented rise in mortality that translated into life expectancy losses around the world, with only a few exceptions. We estimate life expectancy changes in 29 countries since 2020, including most of Europe, the US and Chile, attribute them to mortality changes by age group, and compare them to historic life expectancy shocks. Our results show divergence in mortality impacts of the pandemic in 2021. While countries in Western Europe experienced bounce-backs from life expectancy losses of 2020, Eastern Europe and the US witnessed sustained and substantial life expectancy deficits. Life expectancy deficits among ages 60+ were strongly correlated with measures of vaccination uptake. In contrast to 2020, the age profile of excess mortality in 2021 was younger with those in under-80 age groups contributing more to life expectancy losses. However, even in 2021, registered COVID-19 deaths continued to account for most life expectancy losses.

**Research in context:** *Evidence before this study:* The COVID-19 pandemic disrupted mortality trends around the world. Most high-income countries experienced life expectancy declines in 2020, and emerging evidence from low-to-middle income shows substantial losses in life expectancy with large regional variation. These analyses emphasise the impact of COVID-19 deaths and also highlight the effect of excess mortality at older ages as the main contributor to life expectancy losses, although in some countries working-age mortality also contributed substantially to life expectancy reductions. Among those countries with publicly available data on deaths, only a few, including Denmark, Norway, Finland, Australia, South Korea, Iceland and New Zealand did not experience life expectancy losses.

*Added value of this study:* We estimate life expectancy for 29 countries in 2020-21 and assess bounce-backs versus mounting losses. We determine which age groups contributed most to life expectancy changes in 2021, and analyze how age patterns of excess mortality changed between 2020 and 2021. By projecting pre-pandemic mortality trends into 2020-21 we calculate the life expectancy deficit induced by the pandemic. We compare age-specific life expectancy deficits during fall/winter 2021 against age-specific vaccination uptake by October 1st and further decompose the deficit into contributions from COVID-19 versus non-COVID mortality. To contextualize the magnitude of life expectancy loss, we compare the 2020-21 COVID-19 pandemic with historic mortality shocks over the 20th century. We report results for females, males and the total population.

*Implications of all the available evidence:* Life expectancy is an important summary measure of population health. Over the past decade, improvements in life expectancy have slowed in several countries. The COVID-19 pandemic has compounded these trends and disrupted life expectancy improvements across the globe. The pandemic increased life expectancy inequalities between the 29 low-mortality countries that we analyze, as life expectancy losses were higher among countries with lower pre-pandemic life expectancy. COVID-19 may be a short-term mortality shock, but it is unclear whether countries will bounce back to increasing life expectancy trends in the short- or medium-term. New variants continue the spread of SARS-CoV-2 globally. Variation in preventive measures and vaccine uptake has led to disparate mortality burdens across countries. The long-term mortality impacts of social and economic stressors due to the pandemic are not known, neither are the mortality implications of regular re-infection or Long-COVID. Going forward, it is crucial to understand how and why death rates vary across and within countries.

## Introduction

Period life expectancy (LE) is a summary measure of current population health. If mortality increases in a population, life expectancy declines. Conversely, if mortality declines, life expectancy increases. The measure is age-standardized and thus commonly employed for international comparisons of population health. In this paper we investigate life expectancy changes since the start of the pandemic, distinguishing countries which saw worsening losses from countries which managed to “bounce back” from their life expectancy drop in 2020.

Most countries experienced sizable gains in life expectancy during the second half of the 20th century.^1^ However, at the turn of the 21st century, the rate of improvement in life expectancy slowed down in many high-income countries prior to the COVID-19 pandemic,^2^ such as the USA,^3^ England and Wales,^4^ Scotland,^5^ among others.^3^ The COVID-19 crisis triggered a mortality shock resulting in life expectancy declines in 2020 of a magnitude not observed in the recent history of high-income countries.^6;7;8;9^ While data limitations have precluded in-depth analyses in low-to-middle income countries, emerging evidence suggests even larger losses than those observed in high-income countries, such as in India^10^ and Latin America.^11;12;13^ Only very few countries did not witness declines in life expectancy in 2020, including Norway, Denmark, females in Finland, New Zealand, and Australia.^6;7;14;15^

Fluctuations in life expectancy are not uncommon. Typically, life expectancy declines are quickly followed by bounce backs.^16;17^ In contrast with these short-term fluctuations, however, the COVID-19 pandemic induced global and severe mortality shocks in 2020 and, as of spring 2022, is still ongoing. Throughout 2021, the impact of the pandemic became more heterogeneous across populations with differences in prior infection, non-pharmaceutical interventions, and vaccination uptake, all influencing the pandemic’s course. Emerging estimates of life expectancy losses based on excess deaths suggest that most Western European countries are expected to partly recover from losses observed in 2020, while other countries including the USA and Russia will suffer further life expectancy declines.^18^

We examine life expectancy changes since 2019 in 29 countries, including most of Europe, the USA and Chile using data on all-cause mortality from the Short-Term Fluctuations Database (STMF)^19^ following a previously validated methodology.^6^ We distinguish between annual life expectancy changes and life expectancy deficits, the former showing the raw year to year difference, the later indicating the difference between observed and expected life expectancy had pre-pandemic trends continued. Using decomposition techniques, we describe which age groups, and to what extent registered COVID-19 deaths, contributed to recent trends and deficits in life expectancy. We compare the magnitude and length of the current global life expectancy decline with prominent mortality shocks during the 20th century. All our results are reported for females, males, and the total population. We further investigate associations between life expectancy changes and vaccination uptake. Our results quantify the mortality burden of COVID-19 in 2021 and contribute to the debate about recent trends in life expectancy from a cross-national perspective.

## Results

### Changes in life expectancy since 2019

Among the 29 countries analyzed, 13 countries saw LE bounce-backs from 2020 losses (Austria, Belgium, Switzerland, Spain, Finland, France, England and Wales, Iceland, Italy, Netherlands, Portugal, Sweden, and Slovenia), and 11 witnessed continued (compound) losses (Bulgaria, Chile, Czech Republic, Germany, Estonia, Greece, Croatia, Hungary, Lithuania, Poland, Slovakia). Life expectancy in the USA, Scotland, and Northern Ireland remained at approximately the same depressed levels as 2020, indicating a constant excess mortality. The extremes are marked by Bulgaria, with record compound LE losses across 2020 and 2021, and France, Belgium, Switzerland, and Sweden, all with complete LE bounce-backs from substantial prior losses. Of the three countries that experienced no LE loss in 2020, Denmark, Norway, and Finland, only Norway had a significantly higher LE in 2021 compared to 2019 (Figure 1, Table 1).

**Figure 1:**
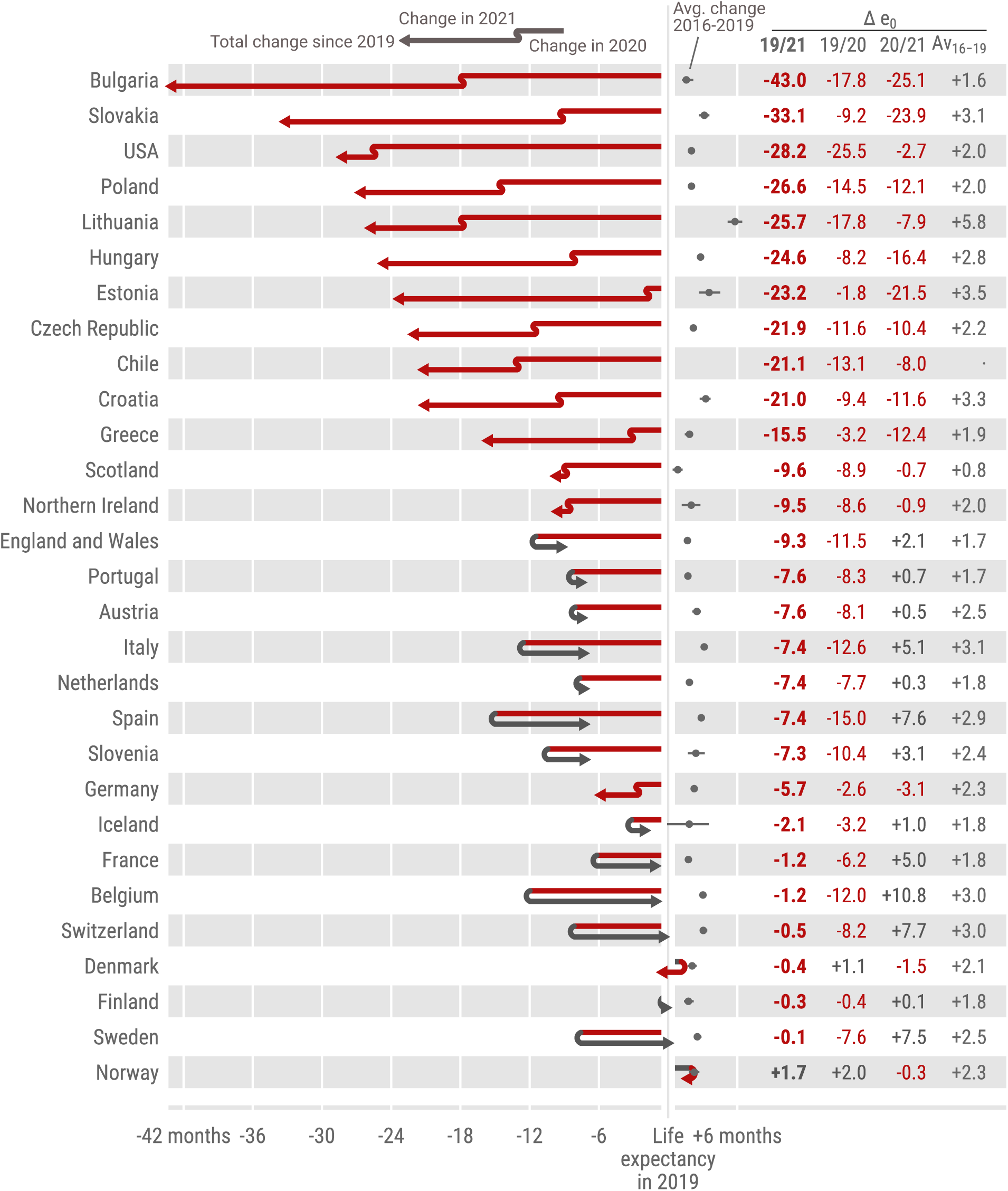
Life expectancy changes 2019-20 and 2020-21 across countries. The countries are ordered by increasing cumulative life expectancy losses since 2019. Grey dots indicate the average annual LE changes over the years 2015 through 2019.

**Table 1:**
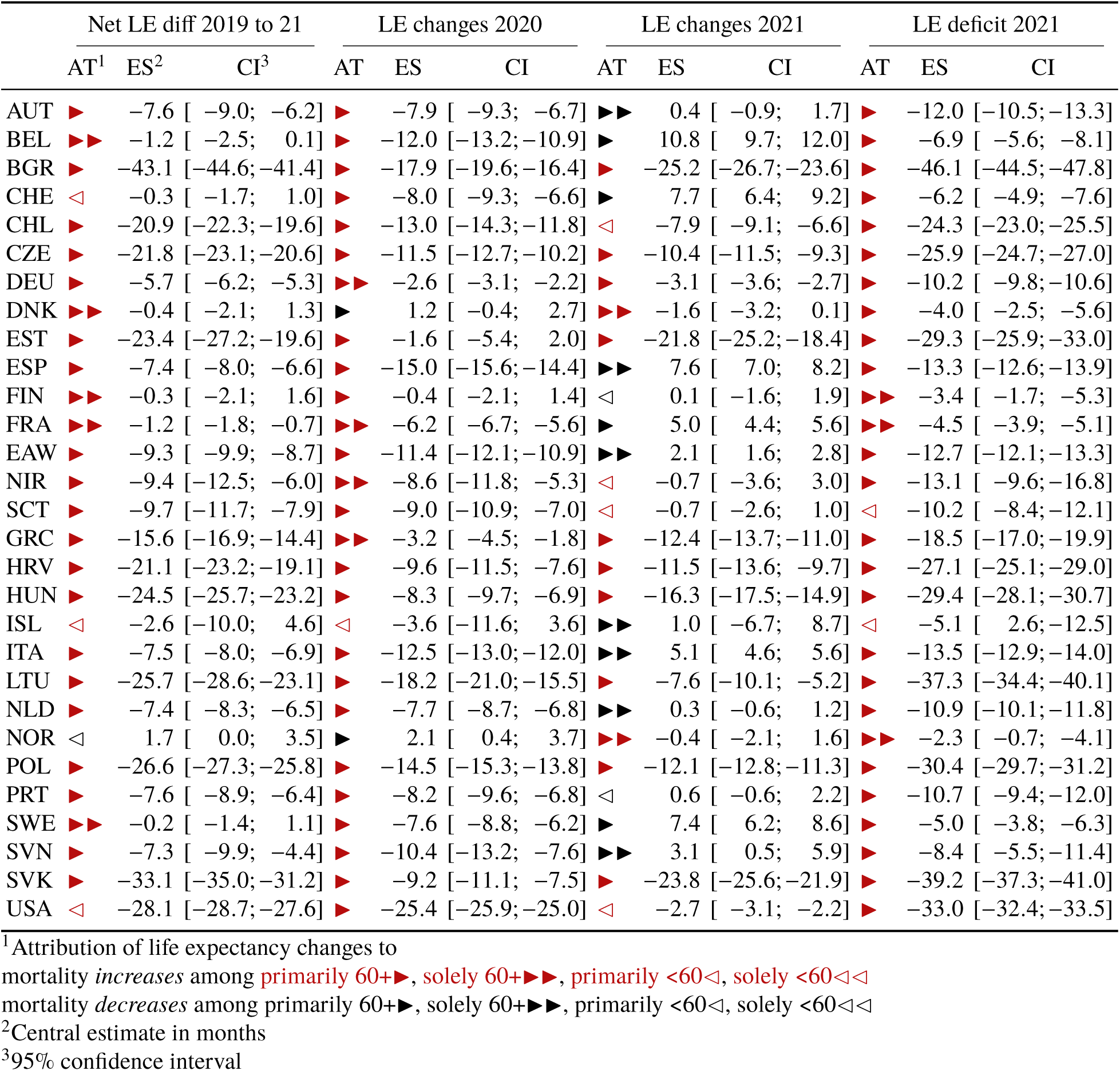
Months of life expectancy (LE) changes and deficits (labelled ES) since the start of the pandemic attributed to age-specific mortality changes (labelled AT). LE deficit is defined as observed minus expected life expectancy had pre-pandemic mortality trends continued.

In all countries LE in 2021 was lower than expected under the continuation of pre-pandemic trends. Bulgaria, Chile, Croatia, Czech Republic, Estonia, Germany, Greece, Hungary, Lithuania, Poland, and Slovakia suffered substantially higher LE deficits in 2021 compared to 2020, indicating a worsening mortality burden over the course of the pandemic (Appendix Figure 12).

Bulgaria experienced 17.9 months of LE decline in 2020 with a 95% CI from 16.4 to 19.6 months. This substantial decline was compounded by an even larger loss of 25.2 (95% CI 23.6, 26.7) months below the 2020 level in 2021, leaving the country with a net LE loss of 43.1 (95% CI 41.4, 44.6) months since 2019. Bulgaria is the most severe example among the 9 countries from the former Eastern Bloc (Bulgaria, Slovakia, Lithuania, Poland, Estonia, Hungary, Czech Republic, Croatia, Slovenia). With the exception of Slovenia, all these countries suffered compound LE losses in 2021. Estonia stands out as the country with the 3rd largest LE losses in 2021 but almost no losses 2020. Substantial compound losses were also observed in Chile and Greece. In contrast, after an 8 month LE loss in 2020, Switzerland experienced a bounce-back of 7.7 months (95% CI 6.4, 9.2). Belgium, Sweden, France, Italy and Spain joined Switzerland as countries that witnessed bounce-backs from substantial LE losses in 2020 with the first 3 countries having regained the LE levels of 2019 (Figure 1, Table 1).

### Age contributions to life expectancy changes

In 2021, the pandemic death toll shifted towards younger age groups. For example, while USA mortality for ages 80+ returned to pre-pandemic levels in 2021, overall LE losses grew due to worsening mortality primarily in ages below 60 (Figure 2). Excess mortality among under-60s explained more than half of the loss in LE in 2021 compared to 2019. LE losses in the under-60s, especially for males, were considerably higher in the USA relative to most other countries in 2020 as well.^6^ This pattern of the shift in excess mortality away from the oldest ages in 2021 compared to 2020 is also evident in Austria, Belgium, Czech Republic, England & Wales, Germany, Netherlands, Northern Ireland, Poland, Portugal, Scotland, Slovakia, Slovenia, and Spain (Figure 2). In 11 out of 16 countries with LE losses in 2021, the under-60 age groups contributed more to LE loss in 2021 than in 2020. Among the 13 countries which partially or completely bounced-back from their LE losses in 2020, 7 (Austria, Spain, England & Wales, Iceland, Italy, Netherlands, Slovenia) achieved the bounce-back solely due to normalizing mortality among the older but not the younger population (Table 1). Croatia, Greece, Hungary, Northern Ireland, and Slovakia saw almost no losses in the 40-59 age group in 2020, but substantial excess mortality in the same group in 2021.

**Figure 2:**
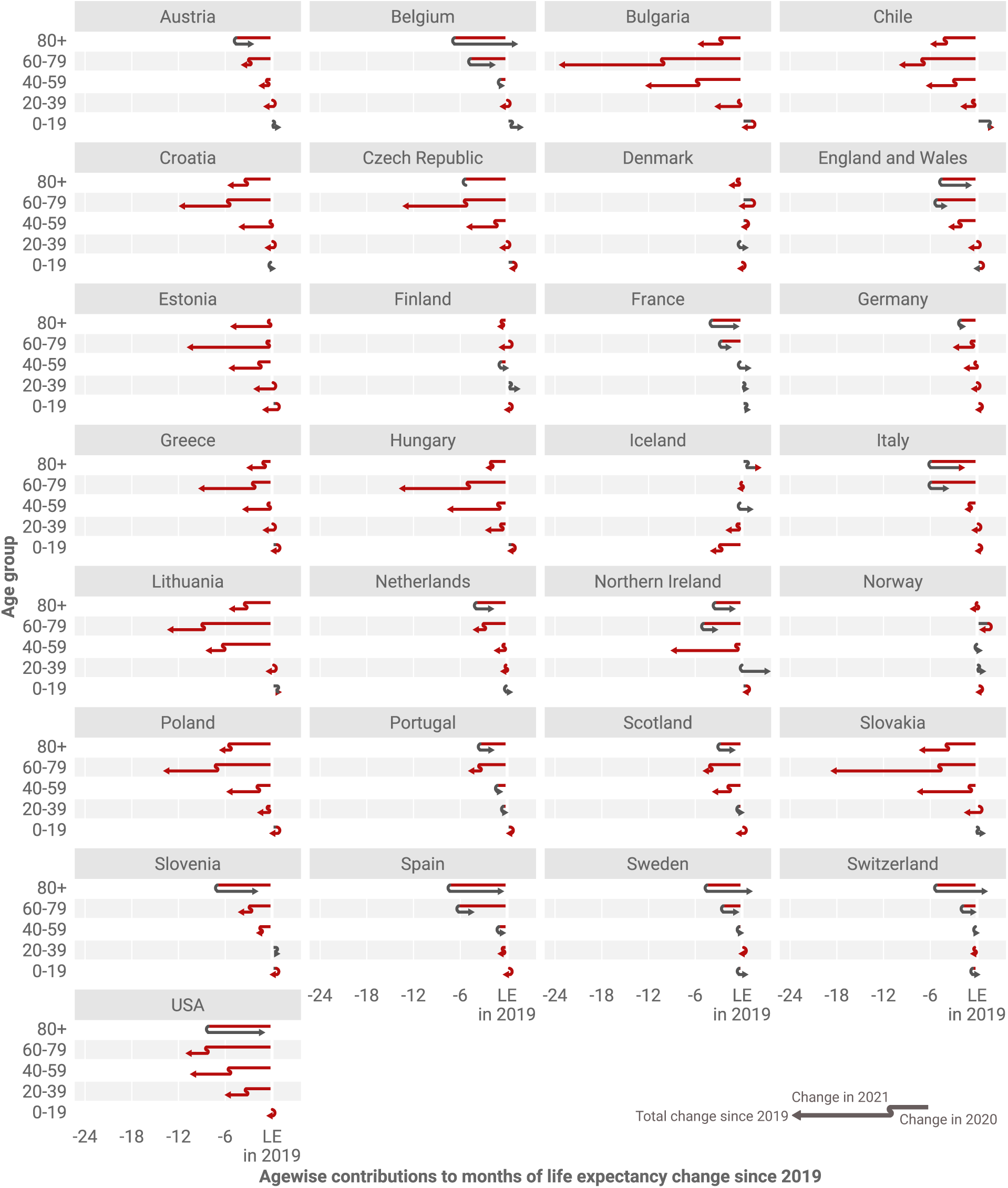
Age contributions to life expectancy changes since 2019 separated for 2020 and 2021. The position of the arrowhead indicates the total contribution of mortality changes in a given age group to the change in life expectancy at birth since 2019. The discontinuity in the arrow indicates those contributions separately for the years 2020 and 2021. Annual contributions can compound or reverse. The total life expectancy change from 2019 to 2021 in a given country is the sum of the arrowhead positions across age.

Despite the shift towards a greater contribution of excess mortality from younger age groups in 2021, increased mortality among those aged 60+ remained the most important contributor to LE losses compared with pre-pandemic levels (Table 1). LE dropped in 28 out of the 29 countries analyzed from 2019 to 2021, with only Norway exceeding the 2019 levels. Excess mortality in ages 60+ was the main contributor to these losses in 25 out of 28 countries, with the USA being the prominent exception. In 2020, the LE losses of every country witnessing significant losses were explained primarily or solely by mortality increases in ages 60+. The LE changes in 2021 were however sometimes driven by mortality dynamics below age 60. France stands out as the only country which suffered substantial life expectancy losses since 2019 without increased mortality among those under-60 (Table 1, Figure 2).

### Sex differences in life expectancy changes

Recent trends of a decreasing gap in LE between females and males^20^ were disrupted by the pandemic. Consistent with previous research, females showed higher life expectancy in the 29 countries in our analysis. However, our results show that the female advantage in LE increased in most of the 29 countries during the pandemic, thereby widening the sex gap (Figure 3). This finding indicates that for most countries males were more affected by excess death. The magnitude of the gap varies from 3 years in the Netherlands to more than 9.7 years in Lithuania. The biggest increase in the sex gap was observed in the USA, where the gap increased by almost a year from 5.72 to 6.69 years. Significant reductions in the LE gap were observed in Bulgaria and Lithuania.

**Figure 3:**
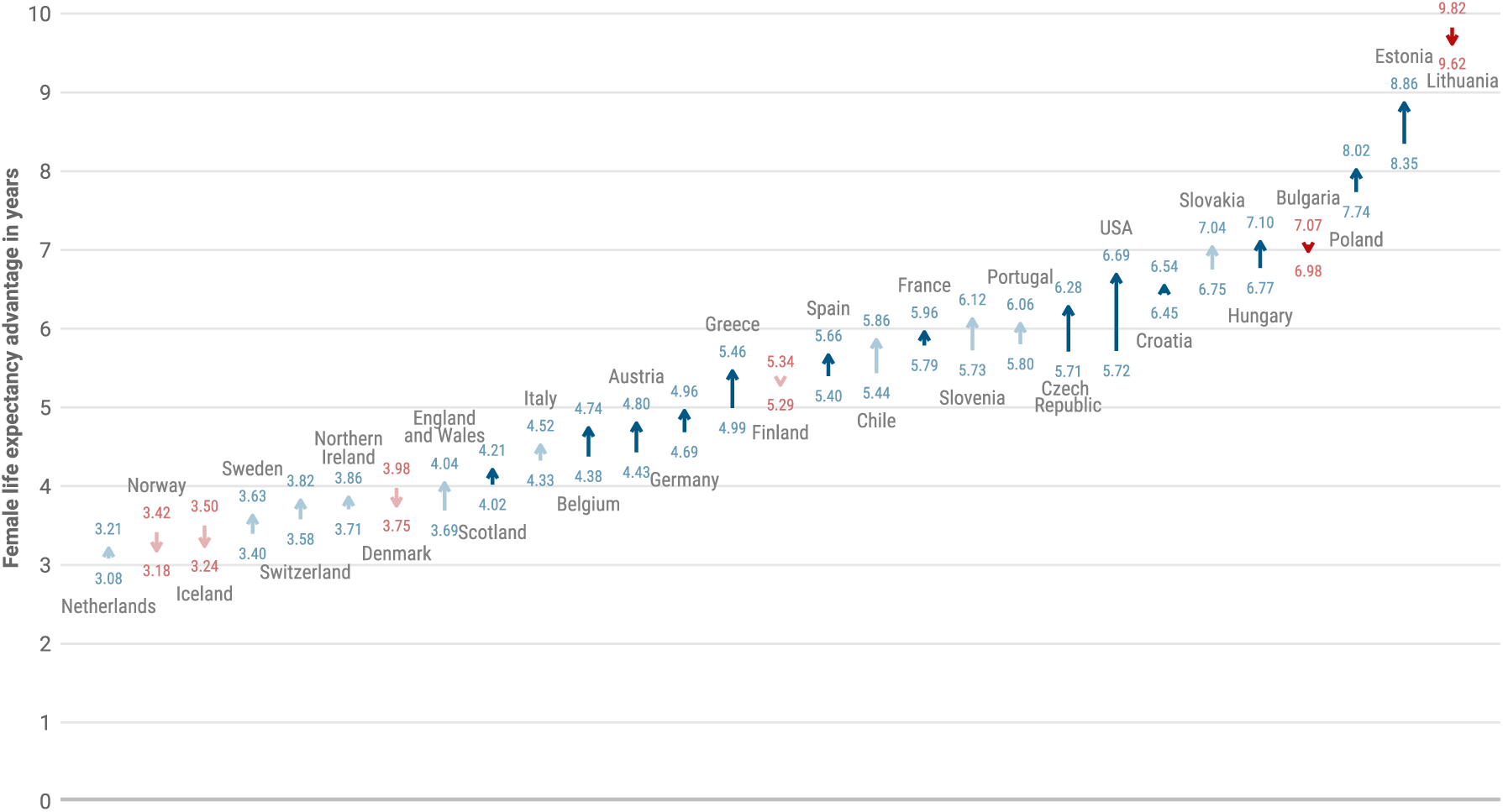
Change in the female life expectancy advantage from 2019 through 2021. Blue colors indicate an increase and red colors a decrease in the female life expectancy advantage. Muted colors indicate non-significant changes.

### Life expectancy deficit contributions by cause of death and age

Officially registered COVID-19 deaths explained most of the LE deficit in the year 2021 across Europe, the USA and Chile (Figure 4). The Netherlands is the single exception where causes other than COVID-19 explained more than half of the 2021 LE deficit. Conversely, France and Slovenia stand out as two countries where mortality due to non-COVID deaths was lower than expected in 2021. In most countries the age group 60–79 contributed the most to the LE deficit in 2021. Exceptions were Scotland and Germany, the former having the largest contributions among ages 40–59, the latter among ages 80+. Note that COVID-19 related deaths are counted differently across countries and that some cross-country differences will be explained by different reporting conventions as outlined in the discussion.

**Figure 4:**
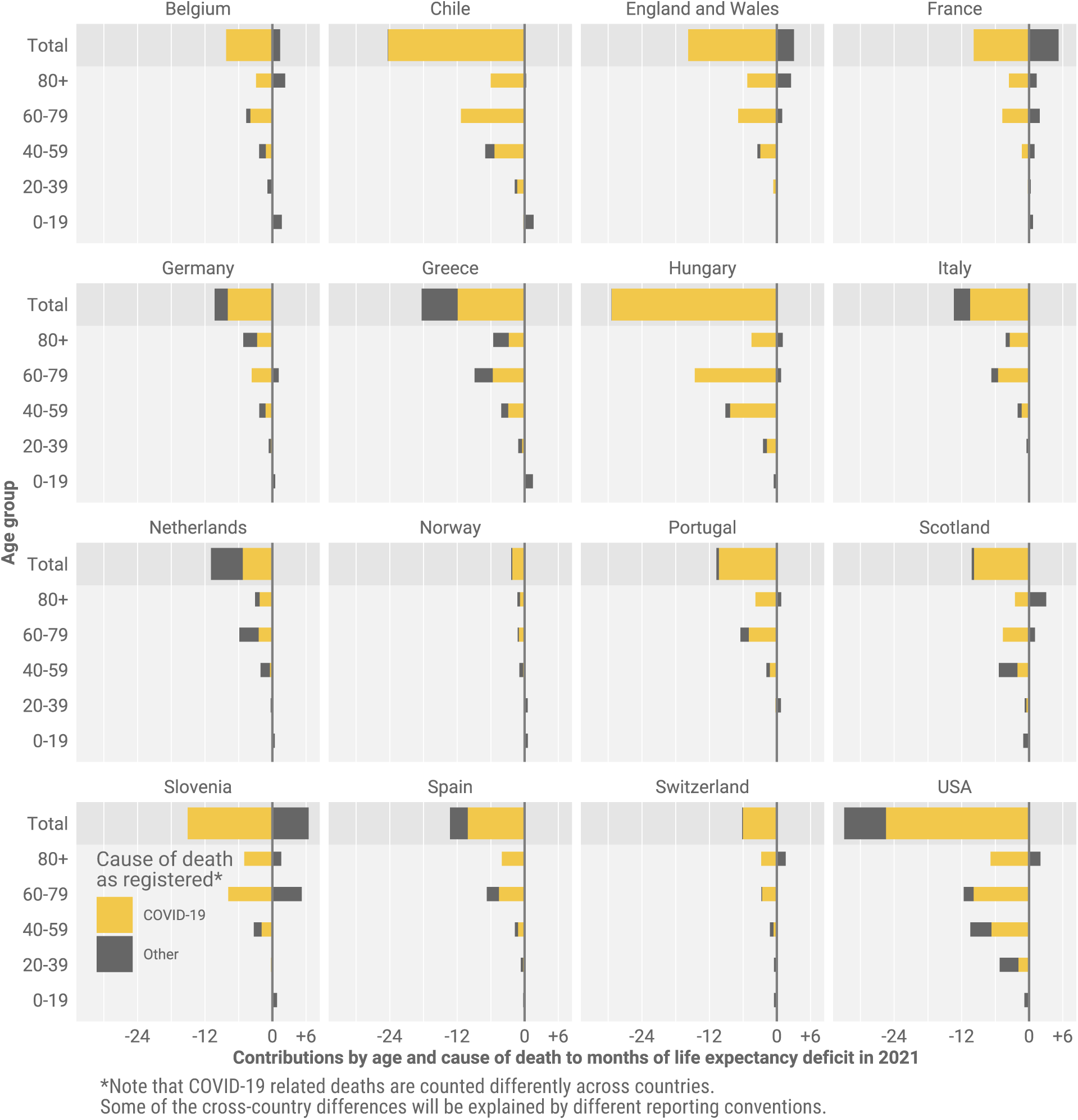
Life expectancy deficit in 2021 decomposed into contributions by age and cause of death. LE deficit is defined as observed minus expected life expectancy had pre-pandemic mortality trends continued.

### Life expectancy deficit by vaccination uptake

Higher vaccination uptake by October 2021 was associated with smaller LE deficits in 2021 across countries (Figure 5). Eastern Europe, especially Bulgaria, which had lower vaccination uptake showed bigger deficits in LE, while the opposite was true for most Central and Western European countries.

**Figure 5:**
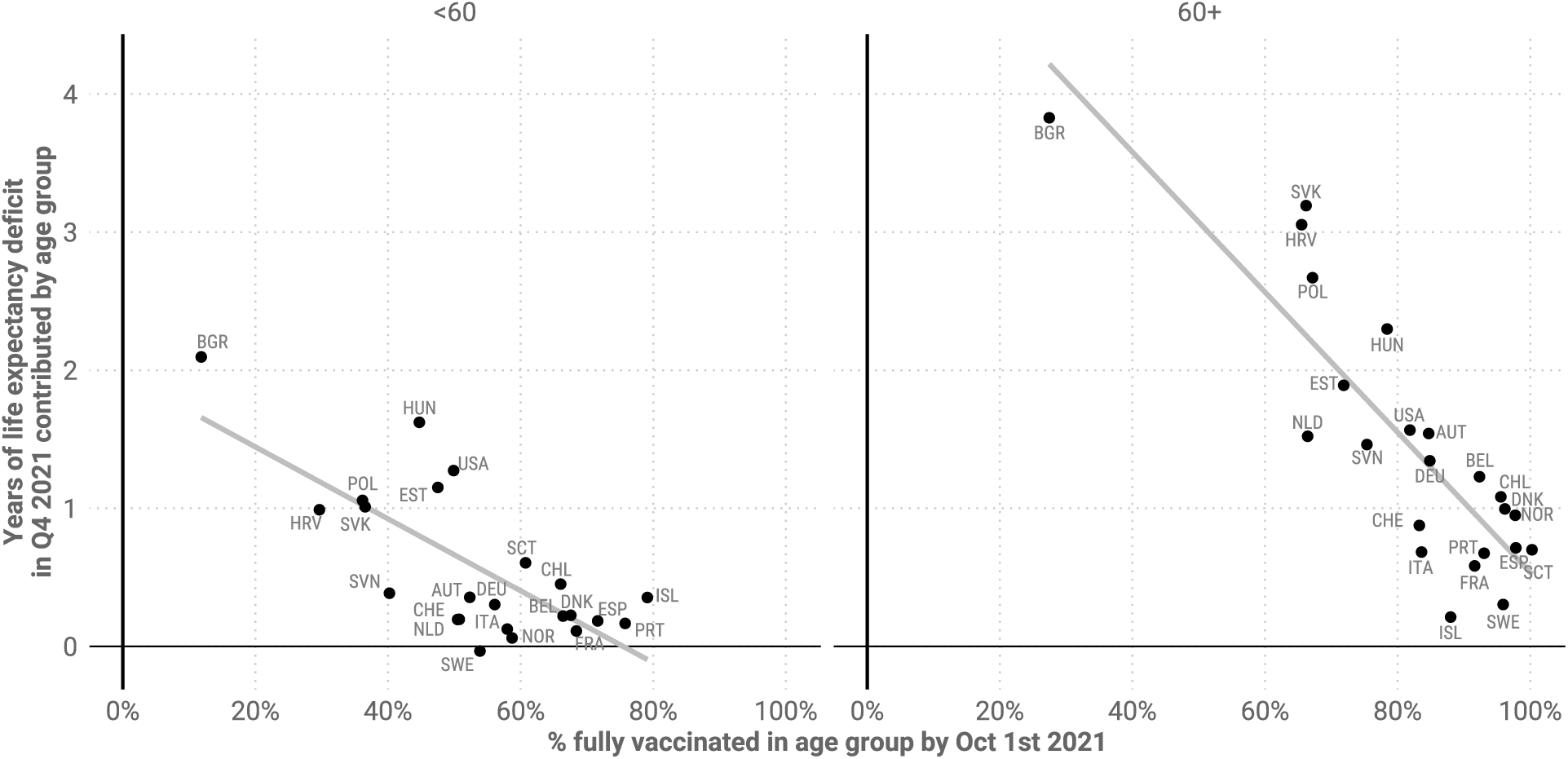
Years of life expectancy deficit during October through December 2021 contributed by ages <60 and 60+ against % of population twice vaccinated by October 1st in the respective age groups. LE deficit is defined as the counterfactual LE from a Lee-Carter mortality forecast based on death rates for the fourth quarter of the years 2015 to 2019 minus observed LE.

The direction of this association was the same when comparing the contributions of the age groups <60 and 60+ to the LE deficit in 2021, albeit with variation in strength of the association. Vaccination uptake for people 60+ showed a stronger association with LE deficits.

Prominent outliers indicate a confounded relationship between vaccination uptake and LE losses. For those under age 60 the USA saw a far higher LE deficit compared to countries with comparable overall vaccination shares, e.g. the Netherlands, Austria, or Switzerland. For ages 60+, Slovakia, Croatia, and Hungary stand out as countries with surprisingly high LE deficits given their vaccination uptake. Finer-grained details of the age prioritization of vaccine roll-out and the types of vaccines used may account for some of these differences, as well as correlations between vaccine uptake and compliance with non-pharmaceutical interventions or the overall health care system capacity.

### Comparison with past mortality shocks

To contextualize the severity of the LE losses during the COVID-19 pandemic, we examined historical mortality crises over the past 120 years and compared them with LE declines since 2019. As shown in Figure 6, the first-half of the 20th century witnessed several mortality shocks leading to LE declines across consecutive years, but in most cases these were followed by immediate bounce-backs. In the past 40 years, the frequency of mortality crises fell markedly. Table 5 in the Appendix shows the total negative or overall change in LE during the period of a mortality crisis and the year at which recovery to pre-crisis LE was reached.

**Figure 6:**
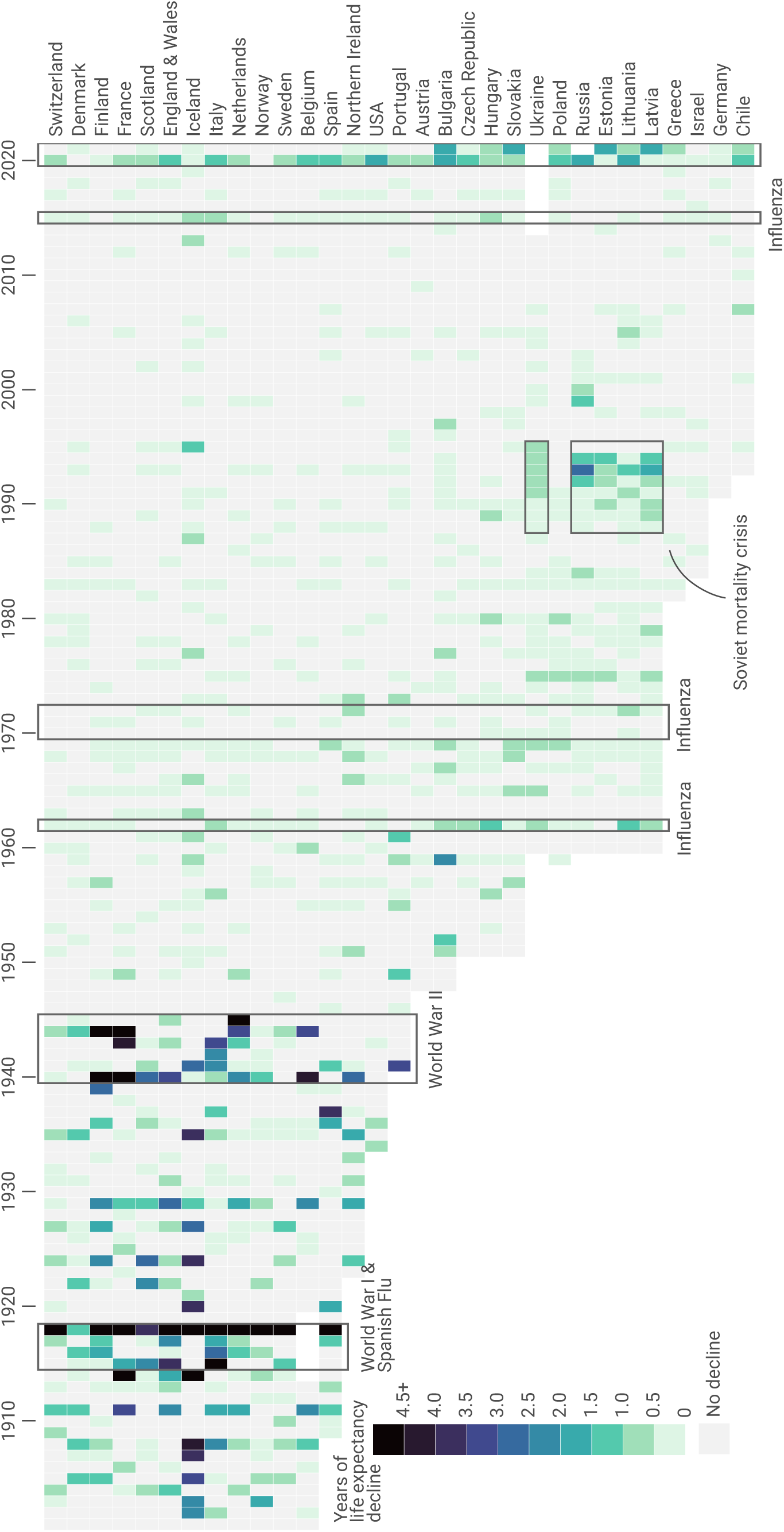
Life expectancy declines since 1900 and periods of widespread mortality increases.

During World War One (WW-I) and the Spanish flu epidemic, all countries for which historical data are available experienced substantial losses in LE – the largest annual declines in LE in the past 120 years. In most countries, LE declined continuously throughout the 4-year period of the crises but the losses were largest in 1918. The steepest declines in LE during 1914–1918 were seen in Italy and France. Denmark, similarly to patterns during COVID-19, experienced the lowest decline in LE during Spanish flu – only 1 year. Notably, even after all these substantial losses to LE, the recovery to pre-crisis levels was achieved in 1 or 2 years (Table 5) .

Mortality patterns during WW-II were somewhat different. While most countries suffered some years with substantial LE declines during 1939-1945, the bounce-backs occurred during the war or just after it. In the Netherlands, the famine known as “Hunger Winter” caused LE declines in 1945 of 5.7 years, but the recovery to pre-war (1938) levels was already by 1946.^21^

Numerous flu epidemics occurred over the second half of the 20th century, but LE during these flu seasons usually declined only very slightly, if at all. Moreover, bounce-backs were always immediate, except in cases of wider health crises, such as the recovery from the 2015 influenza season in Scotland and USA, where LE was already stagnating before the epidemic^5^. Overall, the losses during these flu epidemics were substantially smaller than the declines in LE during the COVID-19 pandemic.

The most prominent example of a protracted mortality crisis in the past 50 years is provided by Russia and Eastern Bloc countries. From 1960s onward, these countries experienced an extended period of continuous stagnation in LE, in which bounce-backs to pre-decline levels were only attained in the 21st century.^22^ These patterns reflect the deep-rooted structural nature of the mortality crisis in the populations that failed to proceed with the health transition.^23^ The most pronounced drop in LE happened in these countries in late 1980s and early 1990s (Figure 6), which mirrored the previous brief period of success in lowering mortality^24^ as a direct result of Gorbachev’s Anti-alcohol campaign.^25^ In contrast to epidemic or war-related shocks, the mortality crisis in the formerly Eastern Bloc countries was structural and the bounce-backs of LE were very slow. The magnitude of LE losses witnessed during COVID-19 in the formerly Eastern Bloc countries are comparable to those observed during the Soviet mortality crisis.

## Discussion

The COVID-19 pandemic led to global mortality increases and declines in period life expectancy which are without precedent over the last 70 years. The scale of these losses was clear by the end of 2020. By the end of 2021, it was clear that the pandemic induced a protracted mortality shock in the USA and many European countries, measured as either compounded LE losses or persistent LE below pre-pandemic levels. Even the best performing countries were lagging behind their life expectancy projections for 2021 given a continuation of pre-pandemic trends. Nevertheless, in 2021 Sweden, Switzerland, Belgium, and France managed to bounce-back from substantial LE losses in 2020 to pre-pandemic levels. Demographically, they achieved this by reducing mortality among ages 60+ back to 2019 levels while avoiding the mortality burden shift to younger ages as seen in other countries during 2021.

Denmark, Finland, and Norway, while lagging behind their projected LE for 2021, managed to remain at pre-pandemic LE levels throughout 2020 and 2021. Here we may see the combination of campaigns delivering vaccines faster to more people than the EU average, effective non-pharmaceutical public health interventions, and high baseline capacities of the health care systems.

It is important to note that even for those countries which approached pre-pandemic levels of life expectancy in 2021, significant damage was already done. This is especially relevant for Sweden which experienced pre-pandemic levels of LE in 2021, but unlike its Scandinavian neighbors, suffered a substantial LE loss in 2020. While period mortality, and consequentially LE can revert back to normal levels, the years of life lost during the period of elevated mortality and declining LE cannot be regained. As a measure of the current mortality conditions in a population, a return to pre-pandemic LE levels simply indicates a normalization of the mortality risk in a population.

We observed stark cross-country differences in 2021 LE deficits with bigger losses in countries with lower pre-pandemic LE. Geographically, this presents as an East-West division in Europe which is also aligned with differences in vaccination uptake during 2021, with generally lower vaccination rates in Eastern Europe compared to the West. This pattern raises questions regarding the future of European mortality convergence, a stated goal of the European Union.^26^

Eastern European countries had a rather distinctive pattern of mortality development in the 20th century. Early on, highly centralized governance and directive economies contributed to mortality reductions through implementation of public health measures, and these countries were very successful in the early stages of their epidemiological transition.^27^ Yet, in the second part of the century these countries witnessed mortality stagnation as these centralized channels were less effective at curbing mortality linked to behavioural factors such as smoking and alcohol.^28;29^ Recent years have seen rapid catch-up life expectancy convergence between Eastern and Western Europe, after the periods of LE declines seen in the 1980s and 90s.^30;23^ It remains to be seen if persistent COVID-19 losses in Eastern Europe compared to diminishing losses in Western Europe will create a new East/West divide and divergence in life expectancy in the years to come.

In the USA the pandemic has accentuated the pre-existing mid-life mortality crisis. This is clear from the strong contribution of increasing mortality below age 60 to LE losses in 2020 and 2021. Because non-COVID mortality also increased in these ages this may be interpreted as the continuation and worsening of a pre-existing mortality crisis among working age adults.^31^ In 2020, the largest share of non-COVID excess deaths in USA males was from external causes (primarily due to drug overdoses and homicides), nearly 80% of which occurred at working ages.^32^ Preliminary data show continued increases in deaths due to drug overdoses in 2021.^33^ However, part of the effect may be due to under-registration of COVID-19 deaths among the working age population. Differences in vaccine uptake by age may also have contributed to the shift to younger mortality in the USA. By July 1st when vaccines where already available in the USA, only 66.9% of 50–64 year-olds were fully vaccinated compared to 82.3% of 65–74 year-olds (as per CDC Covid Data Tracker). This means that older age groups were relatively better protected during the large Delta wave in the USA in the summer/autumn of 2021 compared to previous waves. Pre-pandemic differences in underlying conditions such as obesity and diabetes also may also have contributed to an increased mortality burden in working age USA adults compared to European counterparts.^34^ Regarding global comparisons, the evidence for co-morbidity prevalence as an important predictor of cross-country COVID-19 mortality differences is still weak.^35^

In 2021 the pandemic’s death toll shifted to younger ages. While most countries reduced mortality at older ages, countries that bounced back were able to avoid the mortality increases in those under-60 that accounted for a higher share of losses in 2021. Whether this shift towards the young reflects differences in vaccine protection, behavioral responses, or deaths from indirect causes remains to be understood. The inconsistent registration of deaths due to COVID-19 across countries^36^ complicates any cause-of-death attribution analysis, including ours. We found lower than expected mortality due to non-COVID-19 causes in 2021 in Belgium, England & Wales, France, and Slovenia (Figure 4). If these results are an artefact of an overly broad definition of COVID-19 related deaths or point towards a genuine decline in non-COVID mortality due to e.g. mortality displacement or the lack of flu-deaths is unclear at this point. For France, over-counting COVID-19 deaths seems unlikely as the underlying data on COVID-19 death counts used here originates from Santé Publique France, which uses a very strict definition of “death due to COVID-19”.^36^

COVID-19 was the largest contributor to the 2021 life expectancy deficit in all analyzed countries, but the Netherlands. This is despite different reporting standards and thus provides additional strong evidence for the direct effect of COVID-19 on increases mortality. The result for the Netherlands may be spurious as Karlinsky and Kobak ^37^ found indirect evidence of substantial under-reporting of deaths due to COVID-19 in the Netherlands during the fall/winter wave in 2021 when comparing excess deaths with COVID-19 deaths (we retrieved updated results for 2021 from the authors github repository at github.com/dkobak/excess-mortality on February 15, 2021). However, because factors other than undercounting can influence the relationship between COVID-19 deaths and excess deaths, more detailed cause of death information is needed to assess the impact of non COVID-19 related mortality on 2021 LE deficits.

As the calculation of life expectancy losses requires timely data on population and death counts by age, we are limited in our analysis to those countries with a reliable vital statistics registration system. Consequently our international analysis based on 29 high and middle income countries may give a skewed impression of the global impact of COVID-19 on life expectancy. Indirect life expectancy estimates for 2020-21 based upon excess deaths^18^ indicate substantial losses across South America, which match or exceed the losses we estimated for Eastern Europe. India and select countries in the Middle East likely had losses on par with the U.S. whereas Russia and Mexico suffered life expectancy losses in excess of the 42 months we estimated for Bulgaria. Little is known about the African continent, due to a lack of reliable death registration, or China, due to restrictive access to the data. Putting our results in context with these indirect estimates: while the life expectancy losses in Central, Western and Northern Europe over the first two years of the pandemic have been drastic given the long trend of declining mortality, they likely have been low when compared across the globe.

In 2021 we saw divergence in the impact of the pandemic on population health. While some countries saw bounce-backs from stark life expectancy losses thanks to pharmaceutical and non-pharmaceutical public health interventions, others saw sustained and substantial life expectancy deficits. Human populations faced multiple mortality crises during the 20th century yet life expectancy kept increasing globally in the medium and long term, especially in the second-half of the 20th century. While COVID-19 has been the most severe global mortality shock since the Second World War, we will have to wait to know whether and how longer term life expectancy trends are altered by the pandemic. Extrapolating our findings from 2021 it is plausible that countries with ineffective public health responses will see a protracted health crisis induced by the pandemic with medium-term stalls in life expectancy improvements, while other regions manage a smoother recovery to return to pre-pandemic trends.

## Methods

### LE estimation and decomposition

Calculation of period LE requires data on death counts and population exposures by single years of age. Timely information on death counts across all causes from 2015 through 2021 was sourced from the Short Term Mortality Fluctuations Input Database,^38;19^ which continuously collects weekly death counts by age and sex from statistical offices. Data were downloaded on April 26, 2022. The 38 countries represented in the database have been selected based on completeness of their death registration and census data. To allow for a reliable estimation of life expectancy changes and their age contributions, we further selected those 29 countries for which death counts for 2020 and 2021 were reported across at least 10 distinct age groups. These weekly counts were aggregated into annual counts and, due to various age groupings used in the source data, harmonized into single age groups from 0 to 85+ using the Penalized Composite Link model (PCLM),^39;40^ a non-parametric disaggregation technique for histograms of count data. Separate tables of harmonized counts were created for males, females and the total population in each country. Our methodology for age disaggregation follows that of Aburto et al. (2021)^6^ and has been thoroughly validated for accurate LE estimates consistent with those based on unabridged data. Mid-year population estimates by age and sex for the years 2015–2021 were sourced from the United Nations World Population Prospects (WPP)^41^ and converted into person-years of exposures taking into account the varying number of weeks in an ISO-year^42^ to be consistent with death-counts reporting in our raw data. For Scotland and England and Wales we projected mid-year population for 2020 and 2021 based on data by the respective national statistical offices. Sensitivity analysis with alternative population estimates taken from national statistical offices is provided in the Appendix. In order to attribute changes in LE to changes in mortality from deaths registered as due to COVID-19 we sourced age- and sex-specific COVID-19 death counts from the COVerAGE-DB database.^43^ All data were downloaded on February 18, 2022.

Annual period life tables calculated via standard demographic techniques^44^ were used to estimate LE from 2015 to 2021. To quantify the LE loss directly attributable to COVID-19 we further assembled cause-deleted life tables with “deaths registered as due to COVID-19” and “other deaths” as possible decrements.^45^ Using the Arriaga decomposition technique^46^ we attributed annual changes in LE to changes in age-specific all-cause and cause-specific mortality. Additionally, we calculated LE deficits for 2020 and 2021 defined as observed LE minus expected LE based on a continuation of pre-pandemic trends. These expected LE were derived from Lee-Carter forecasts^47^ of age-specific death rates over the years 2015 through 2019. Confidence intervals around our LE estimates, LE differences, LE deficits, and LE decompositions were derived from 500 Poisson simulations of the harmonized death counts.

To analyze the cross-country relationship between life expectancy deficits and vaccination uptake during fall and winter 2021 we calculated life tables and life expectancy deficits for the fourth quarter of the year. To do so we used the weekly death counts over weeks 40 through 52 and adjusted population exposures.

The analyses are fully reproducible with source code and data archived with Zenodo (DOI: 10.5281/zenodo.6653291). Additionally we archived CSV files of the table and figure data (DOI: 10.5281/zenodo.6653120), and the harmonized life table data informing all analyses in this paper (DOI: 10.5281/zenodo.6653179).

### Estimation of age specific vaccination uptake

Data on age-specific vaccinations was collected from the COVerAGE-DB.^43^ Vaccination uptake was calculated with midyear population data for 2020 from WPP^41^ and the Human Mortality Database. Vaccination measures were age harmonized using the PCLM. Vaccination counts with missing age information were redistributed according to the observed age distribution of vaccinations in a given country. From the available data, for each country we calculated the share of population fully vaccinated as of October 1st 2021 separately for people below age 60 and above 60. A full vaccination was defined as either two vaccinations or a single vaccination of the vaccine from Johnson & Johnson.

## Data availability

We archived CSV files of the table and figure data (DOI: 10.5281/zenodo.6653120), and the harmonized life table data informing all analyses in this paper (DOI: 10.5281/zenodo.6653179).

## Code availability

Scripts for the R programming language to download the source data for our analysis and to reproduce the results in this paper were archived with Zenodo (DOI: 10.5281/zenodo.6653291).

## Data Availability

The analyses are fully reproducible with source code and data archived with Zenodo (DOI:10.5281/zenodo.6653291). Additionally we archived CSV files of the table and figure data (DOI:10.5281/zenodo.6653120), and the harmonized life table data informing all analyses in this paper
(DOI:10.5281/zenodo.6653179).

https://zenodo.org/record/6653291

https://zenodo.org/record/6653120

https://zenodo.org/record/6653179

## Acknowledgements

The authors like to thank Ariel Karlinsky for his help in determining the death count late registration quotas via his invaluable work on the World Mortality Database.

The authors acknowledge the following funding: European Union Horizon 2020 research and innovation programme under the Marie Sklodowska-Curie grant agreement No 896821(J.M.A., R.K.); ROCKWOOL Foundation’s Excess Deaths grant (J.M.A., I.K.); Leverhulme Trust Large Centre Grant (J.M.A., L.Z., R.K., J.B.D.); European Research Council grant ERC-2021-CoG-101002587 (MORTAL) (J.B.D.); University of Oxford John Fell Fund (J.M.A., L.Z., R.K., J.B.D.); Estonian Research Council grant PSG 669 (H.J.)

## Author contributions

Conceptualization (J.S., J.M.A., I.K., M.S.K., H.J., R.K.); Data curation (J.S., M.S.K., I.K., L.Z., H.J., R.K.); Formal analysis (J.S., I.K., M.S.K., L.Z., H.J., R.K.); Investigation (J.S., J.M.A., I.K., M.S.K., L.Z., H.J, J.B.D., R.K.); Methodology (J.S., J.M.A., M.S.K., L.Z., R.K.); Project administration (J.S.); Software (J.S., M.S.K., I.K., H.J.); Supervision (J.S., J.M.A., R.K.); Validation (J.S., J.M.A., I.K., M.S.K., L.Z., H.J., J.B.D., R.K.); Visualization (J.S., I.K., H.J.); Writing – original draft (J.S., J.M.A., I.K., M.S.K., L.Z., H.J., J.B.D., R.K.); Writing – review & editing (J.S., J.M.A., I.K., M.S.K., L.Z., H.J., J.B.D., R.K.).

## Appendix

### Population exposures sensitivity analysis

The results presented in the main text rely on population exposures from the UN World Population Prospects (WPP), which were issued prior to the COVID-19 pandemic. The COVID-19 pandemic has altered population age structure in some countries, reducing the number of people ages 60 and older. The WPP population projections used in this paper do not take this effect into account and thus may overestimate the population in the oldest age groups in the year 2020 and 2021 resulting in negatively biased death rates and positively biased bounce backs. To test how sensitive our results are to the WPP-based denominators, we performed a sensitivity analysis to compare WPP-based denominators used in the analysis with population estimates (or projections in most cases for 2021) reported by the national statistical offices (NSOs) of the countries analyzed. We then assessed the effects of using these two different sources of population exposures on life expectancy estimates.

As shown in Table 2, the overall differences between WPP and NSO mid-year population estimates for 2019, 2020 and 2021 (when available) are generally small and are not consistently positive or negative. Countries that show the largest deviations (e.g. France) in fact show positive deviations, i.e. WPP-based population estimates are smaller than those reported for national statistical offices for 2020. In further, age-specific comparisons, we find a high correlation between the WPP and NSO-based population estimates.

**Table 2:** Deviation of overall midyear population estimates (in 10,000) between UN World Population Prospect (WPP) and National Statistical Office (NSO) estimates.

|  | 2019 Population |  |  | 2020 Population |  |  | 2021 Population |  |  |
| --- | --- | --- | --- | --- | --- | --- | --- | --- | --- |
|  | WPP | NSO | Dif<sup>1</sup> | WPP | NSO | Dif<sup>1</sup> | WPP | NSO | Dif<sup>1</sup> |
| AUT | 895.5 | 887.9 | -7.6 | 900.6 | 891.7 | -9.0 | 904.3 | 896.1 | -8.2 |
| BEL | 1153.9 | 1146.2 | -7.7 | 1159.0 | 1150.7 | -8.3 |  |  |  |
| BGR | 700.0 | 697.6 | -2.4 | 694.8 | 693.4 | -1.4 |  |  |  |
| CHE | 859.1 | 857.5 | -1.6 | 865.5 | 863.8 | -1.6 | 871.5 | 871.6 | 0.0 |
| CHL | 1895.2 | 1910.7 | 15.5 | 1911.6 | 1945.8 | 34.2 | 1921.2 | 1967.8 | 46.6 |
| CZE | 1068.9 | 1067.2 | -1.7 | 1070.9 | 1069.8 | -1.1 |  |  |  |
| DEU | 8351.7 | 8309.3 | -42.4 | 8378.4 | 8316.1 | -62.3 | 8390.0 | 8331.9 | -58.2 |
| DNK | 577.2 | 581.4 | 4.3 | 579.2 | 582.5 | 3.3 | 581.3 | 585.0 | 3.7 |
| EAW | 5924.6 | 5944.0 | 19.4 | 5937.0 | 5972.0 | 35.0 | 5947.9 | 5998.0 | 50.1 |
| ESP | 4673.7 | 4710.5 | 36.9 | 4675.5 | 4735.6 | 60.1 | 4674.5 | 4732.7 | 58.1 |
| EST | 132.6 | 132.7 | 0.1 | 132.7 | 132.9 | 0.3 | 132.5 | 132.6 | 0.1 |
| FIN | 553.2 | 552.2 | -1.1 | 554.1 | 553.0 | -1.1 | 554.8 | 554.0 | -0.8 |
| FRA | 6513.0 | 6721.6 | 208.6 | 6527.4 | 6734.7 | 207.4 |  |  |  |
| GRC | 1047.3 | 1072.2 | 24.8 | 1042.3 | 1161.8 | 119.5 |  |  |  |
| HRV | 413.0 | 406.5 | -6.5 | 410.5 | 404.8 | -5.8 |  |  |  |
| HUN | 968.5 | 977.1 | 8.6 | 966.0 | 975.0 | 9.0 |  |  |  |
| ISL | 33.9 | 36.1 | 2.2 | 34.1 | 36.6 | 2.5 | 34.3 | 37.3 | 3.0 |
| ITA | 6055.0 | 5972.9 | -82.1 | 6046.2 | 5943.9 | -102.3 | 6036.7 | 5916.1 | -120.7 |
| LTU | 276.0 | 279.4 | 3.5 | 272.2 | 279.5 | 7.3 | 269.0 | 278.7 | 9.7 |
| NIR | 189.0 | 189.4 | 0.4 | 189.7 | 189.6 | -0.1 | 190.3 | 190.2 | -0.2 |
| NLD | 1709.7 | 1734.5 | 24.8 | 1713.5 | 1744.2 | 30.7 |  |  |  |
| NOR | 537.9 | 534.8 | -3.1 | 542.1 | 538.0 | -4.2 | 546.6 | 540.5 | -6.1 |
| POL | 3788.8 | 3838.6 | 49.9 | 3784.7 | 3835.4 | 50.8 | 3779.7 | 3816.2 | 36.5 |
| PRT | 1022.6 | 1028.6 | 6.0 | 1019.7 | 1029.7 | 10.0 |  |  |  |
| SCO | 543.7 | 546.3 | 2.6 | 543.1 | 546.6 | 3.5 | 542.4 | 546.9 | 4.6 |
| SVK | 545.7 | 545.4 | -0.3 | 546.0 | 545.9 | -0.1 |  |  |  |
| SVN | 207.9 | 208.9 | 1.1 | 207.9 | 210.0 | 2.1 | 207.9 | 210.7 | 2.8 |
| SWE | 1003.6 | 1027.9 | 24.2 | 1009.9 | 1035.3 | 25.4 | 1016.0 | 1040.4 | 24.4 |
| USA | 32906.5 | 32833.0 | -73.5 | 33100.3 | 32948.4 | -151.9 | 33291.5 | 33499.8 | 208.3 |

<sup>1</sup>Differences between the WPP and NSO midyear population estimates.

Figure 7 shows estimates of life expectancy (LE) using NSO-based denominators compared to WPP-based denominators for 2019, 2020 and 2021 when available. Life expectancy estimates using the two different sources for denominators are concordant and largely lie on or close to the x=y line. When the two diverge, the divergence shows NSO-based LE estimates slightly overestimating LE relative to WPP estimates in 2019. While the direction of the LE changes across both sources is thus the same, the more optimistic LE levels in 2019 imply that NSO-based denominators indicate slightly larger losses in LE between 2019 and 2020, and for the few countries for which 2021 are available, more positive bounce-backs in 2021.

**Figure 7:**
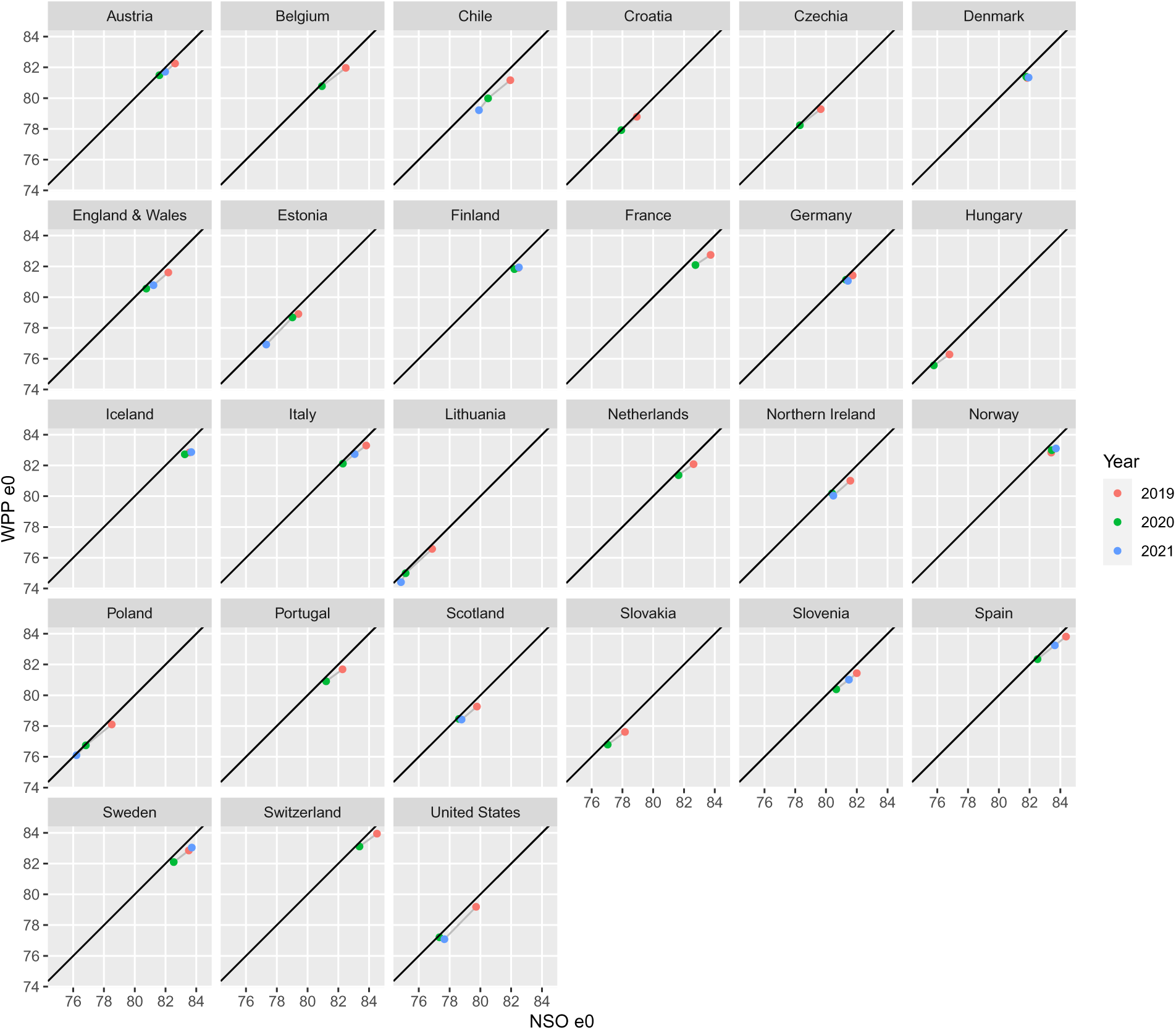
Life expectancy (e0) estimates for 2019, 2020 and when available 2021, using population estimates from national statistical offices (NSOs) (x-axis) and UN World Population Prospects (WPP (y-axis). Black line indicates x=y line.

### Age attributed life expectancy losses and deficits by sex

**Table 3:**
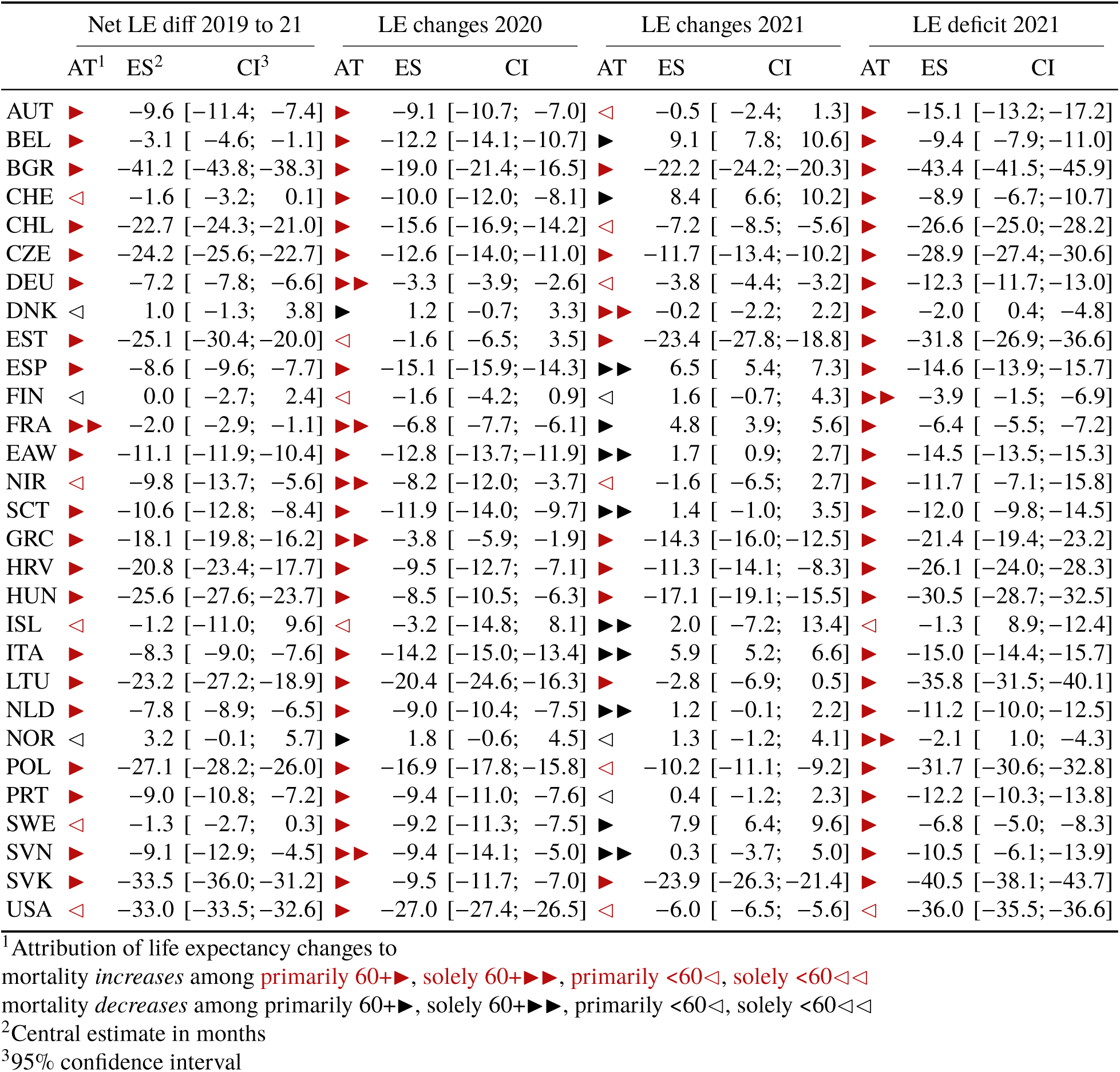
Months of male life expectancy (LE) changes and deficits (labelled ES) since the start of the pandemic attributed to age-specific mortality changes (labelled AT). LE deficit is defined as observed minus expected life expectancy had pre-pandemic mortality trends continued.

**Table 4:**
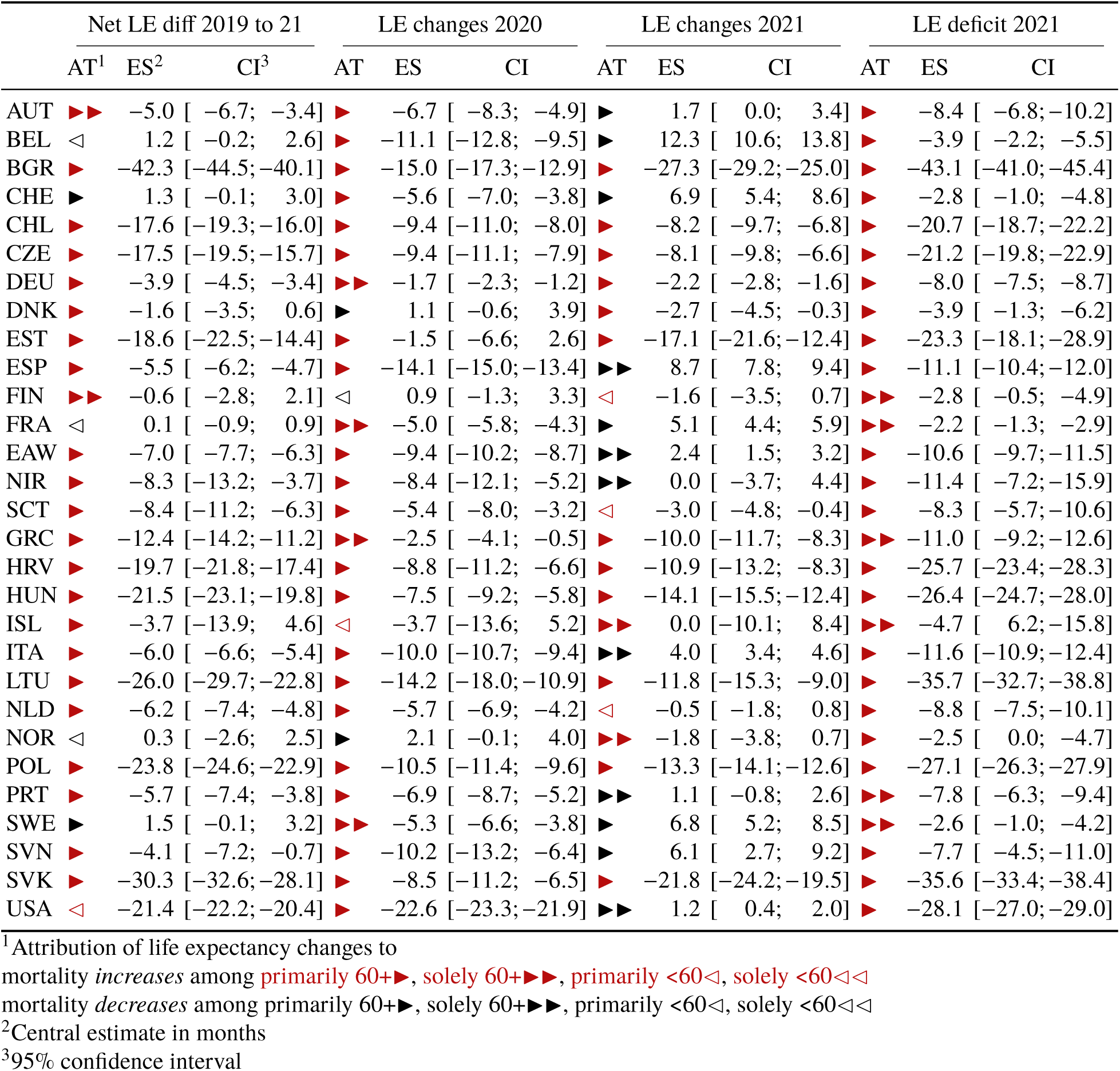
Months of female life expectancy (LE) changes and deficits (labelled ES) since the start of the pandemic attributed to age-specific mortality changes (labelled AT). LE deficit is defined as observed minus expected life expectancy had pre-pandemic mortality trends continued.

### Life expectancy changes since 2019 by sex

**Figure 8:**
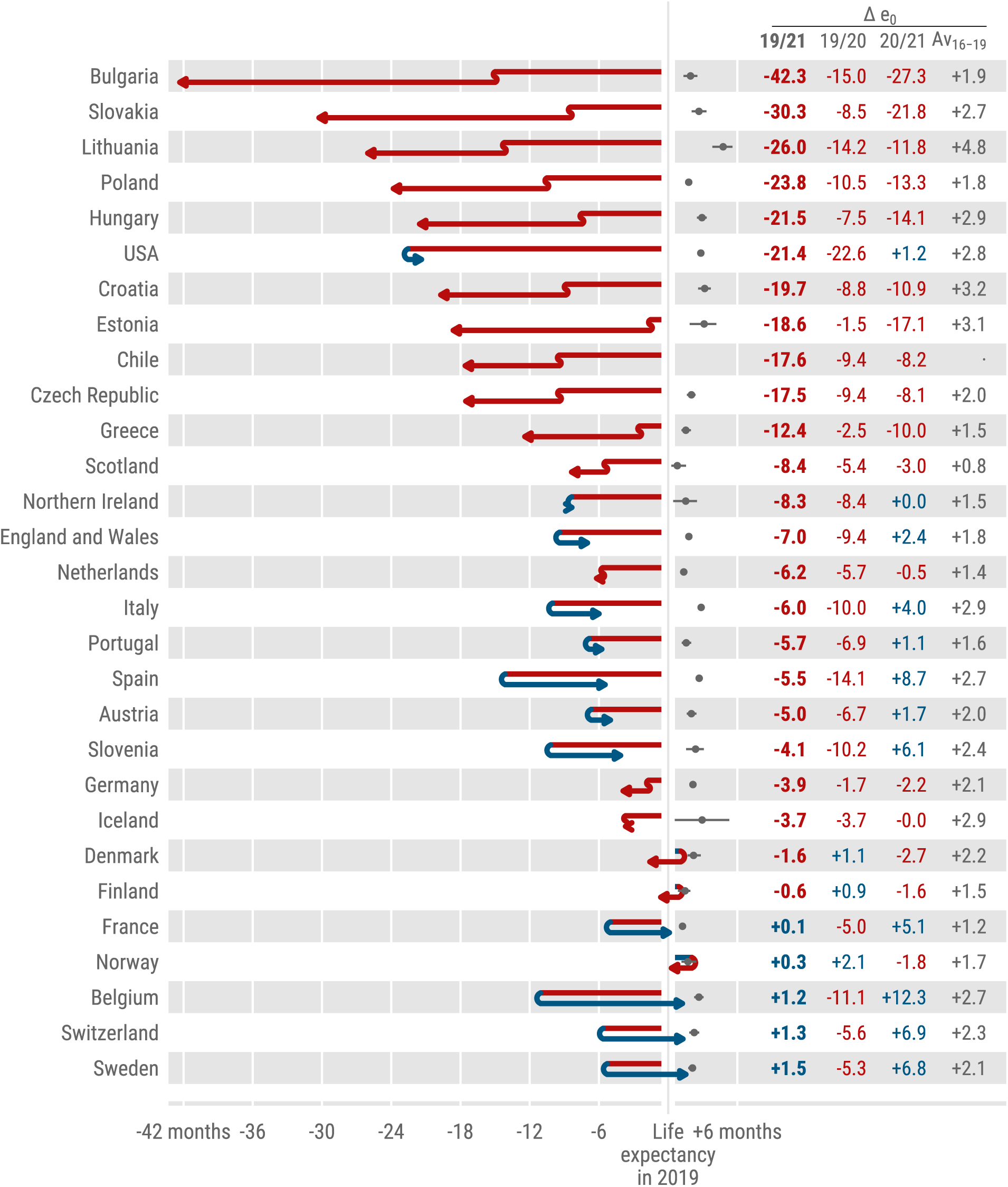
Female life expectancy changes 2019-20 and 2020-21 across countries. The countries are ordered by increasing cumulative life expectancy losses since 2019. Grey dots indicate the average annual LE changes over the years 2015 through 2019.

**Figure 9:**
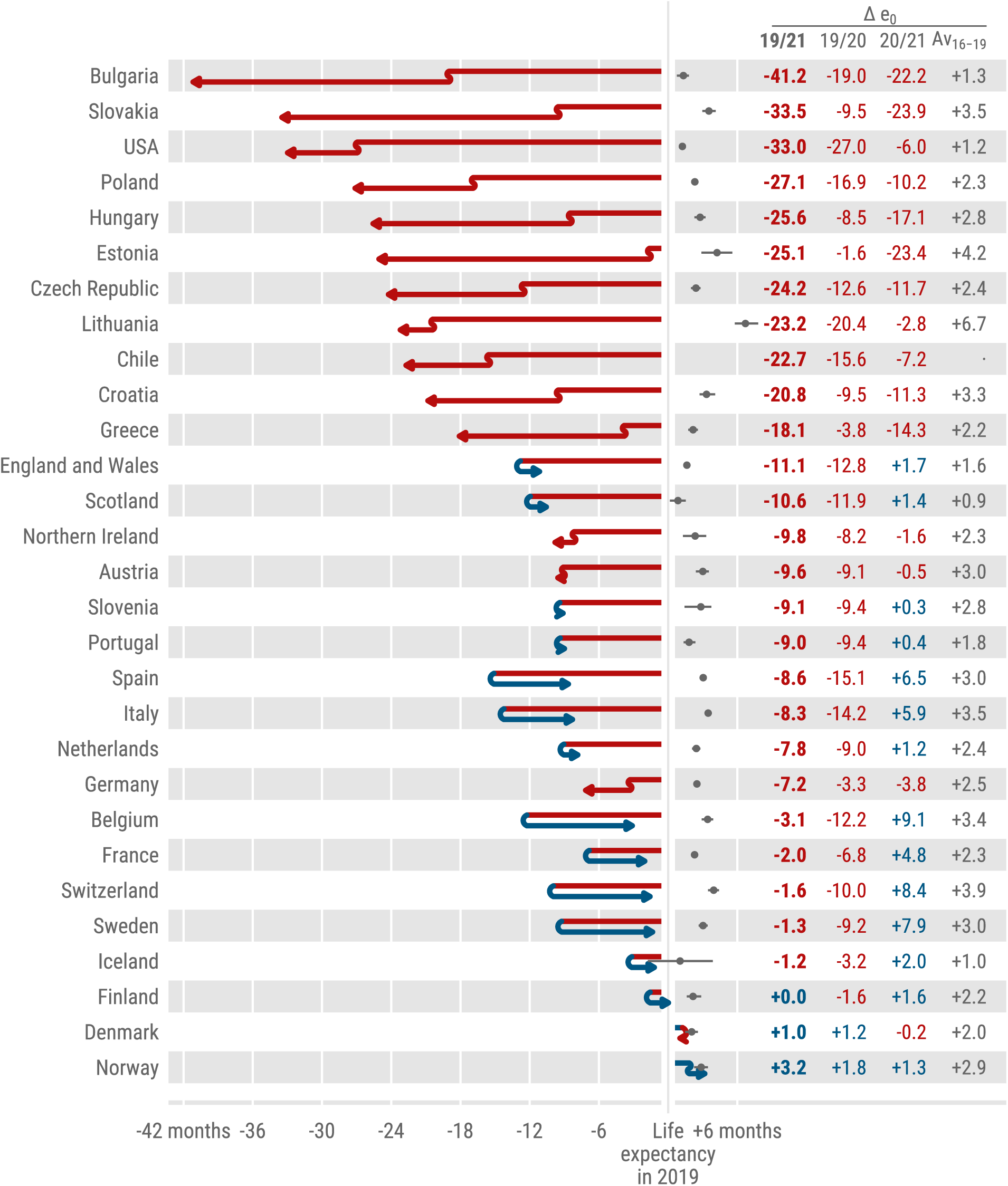
Male life expectancy changes 2019-20 and 2020-21 across countries. The countries are ordered by increasing cumulative life expectancy losses since 2019. Grey dots indicate the average annual LE changes over the years 2015 through 2019.

### Pre-pandemic life expectancy forecasts for 2020 and 2021 by sex

**Figure 10:**
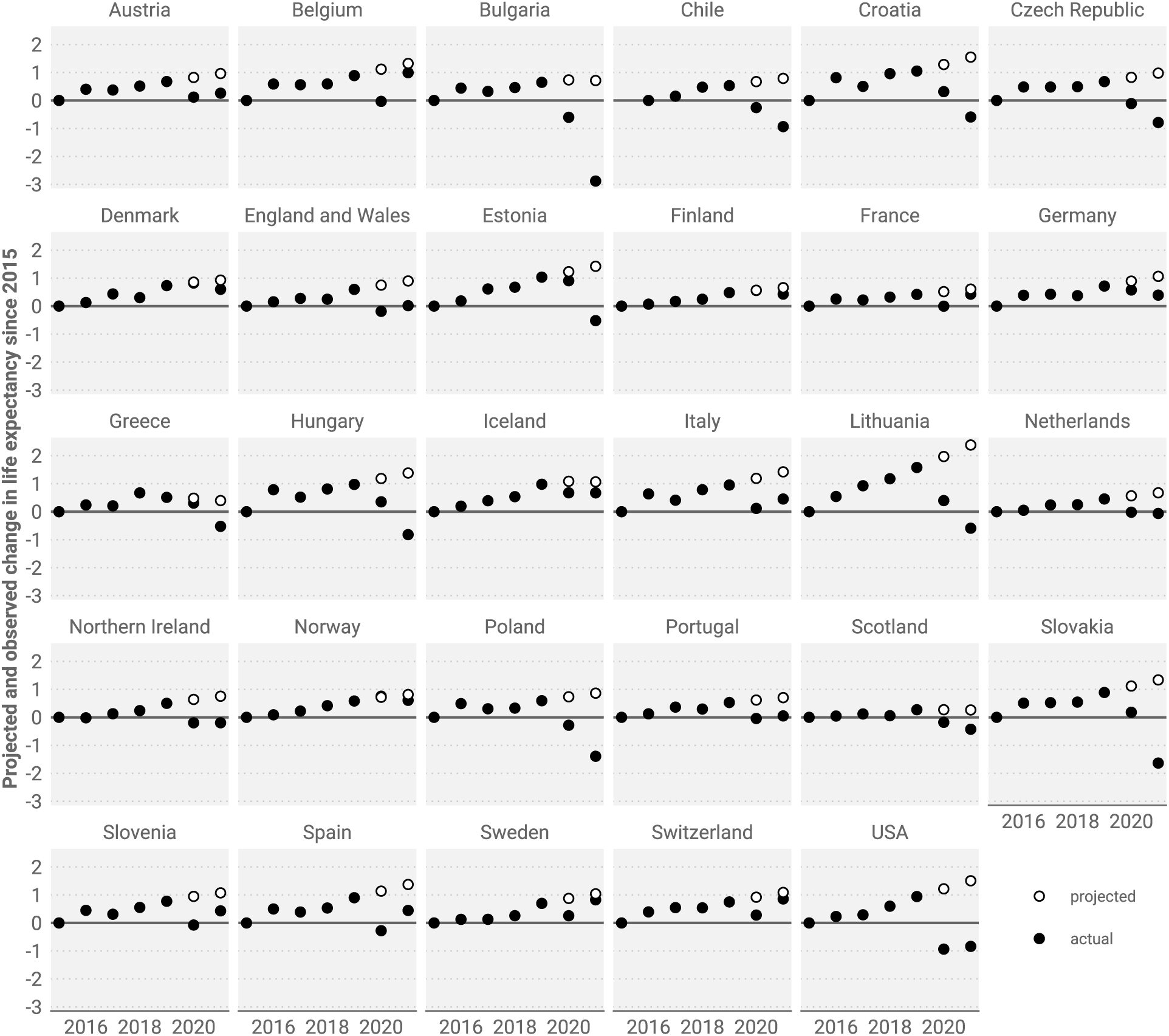
Actual and forecast female life expectancy change since 2015. LE forecasts are based the Lee-Carter model based on the assumption that pre-pandemic mortality trends would have continued into 2020 and 2021.

**Figure 11:**
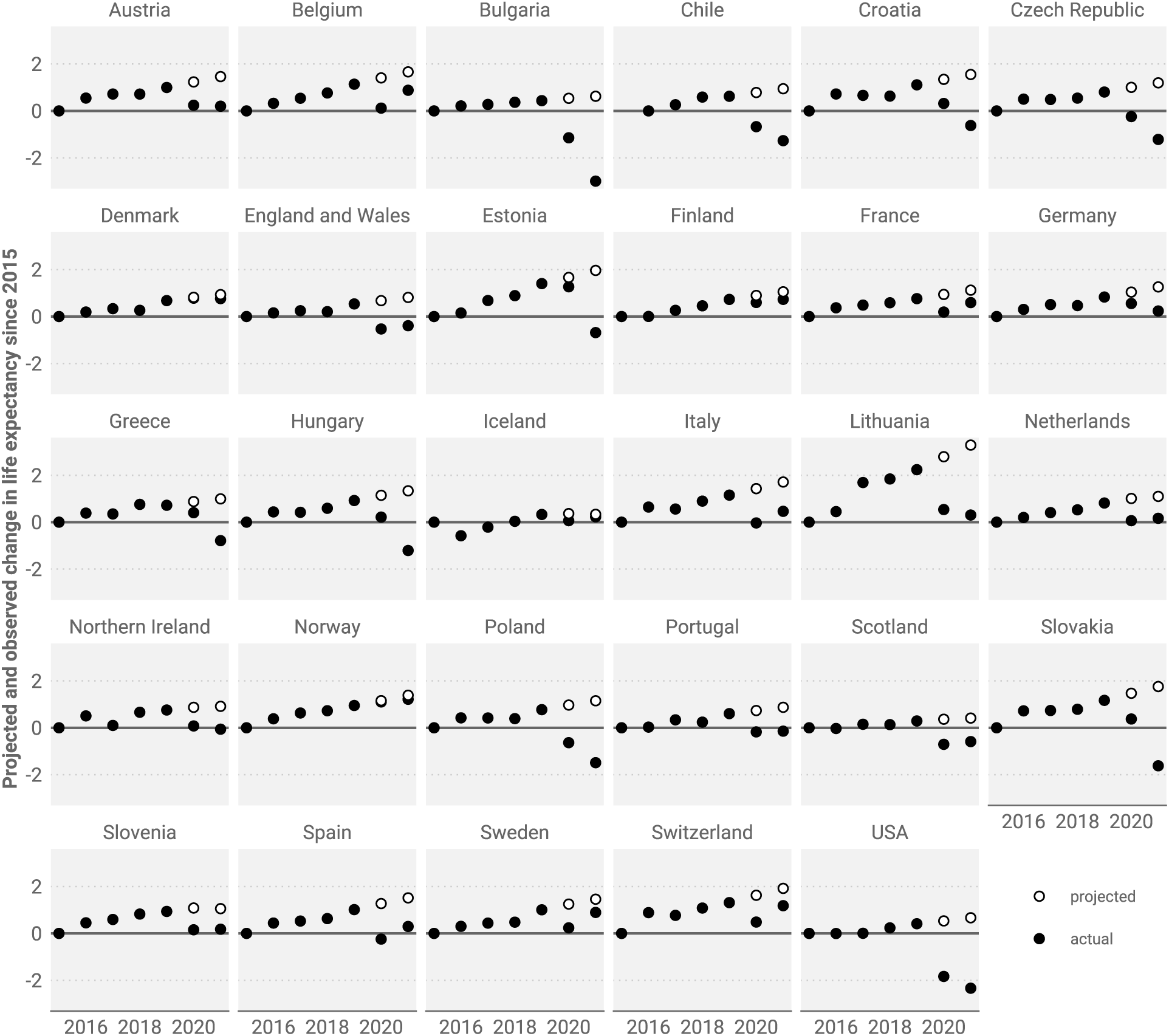
Actual and forecast female male expectancy change since 2015. LE forecasts are based the Lee-Carter model based on the assumption that pre-pandemic mortality trends would have continued into 2020 and 2021.

**Figure 12:**
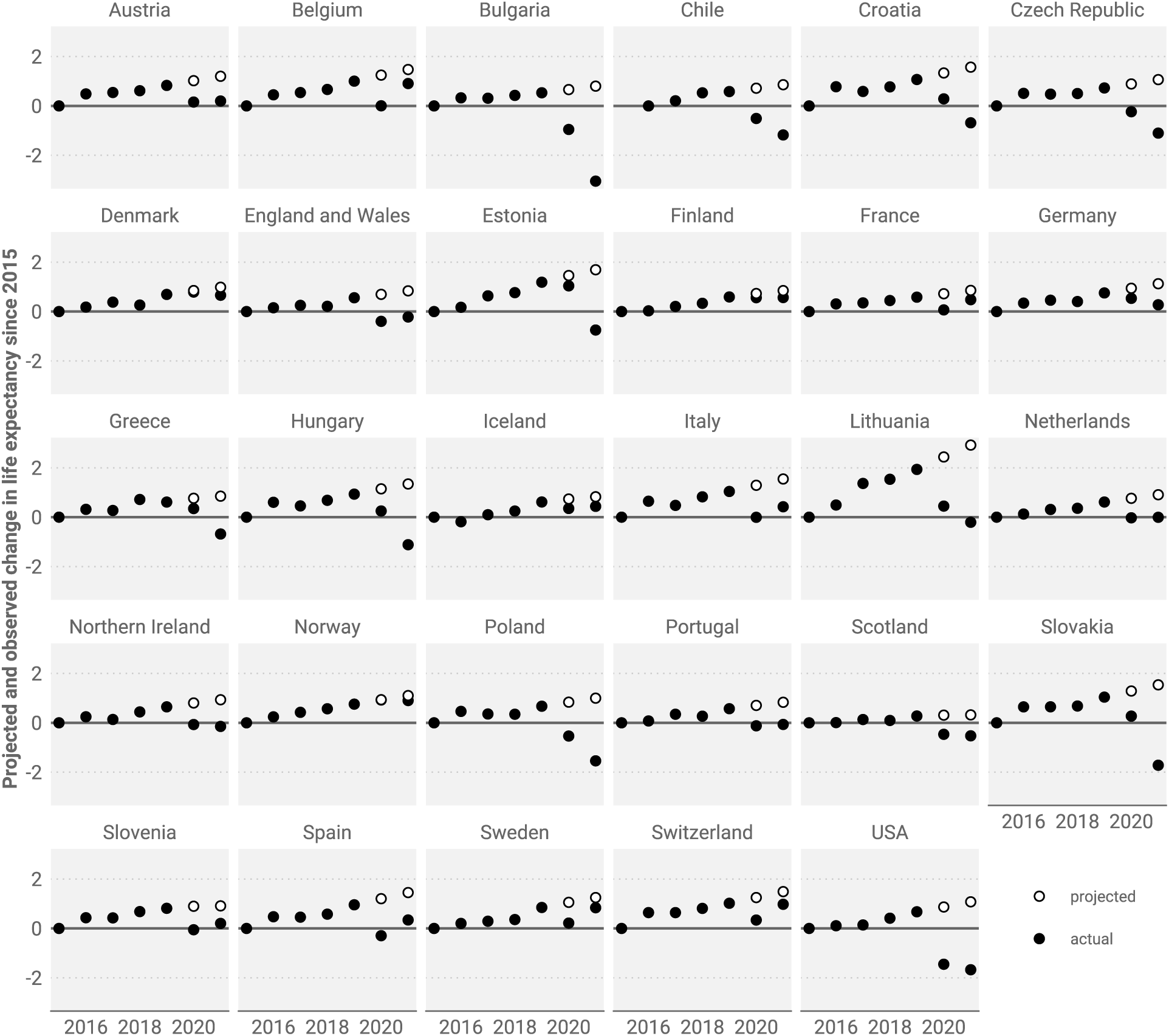
Actual and forecast total population life expectancy change since 2015. LE forecasts are based the Lee-Carter model based on the assumption that pre-pandemic mortality trends would have continued into 2020 and 2021.

### Cause of death contributions to life expectancy deficit by sex

**Figure 13:**
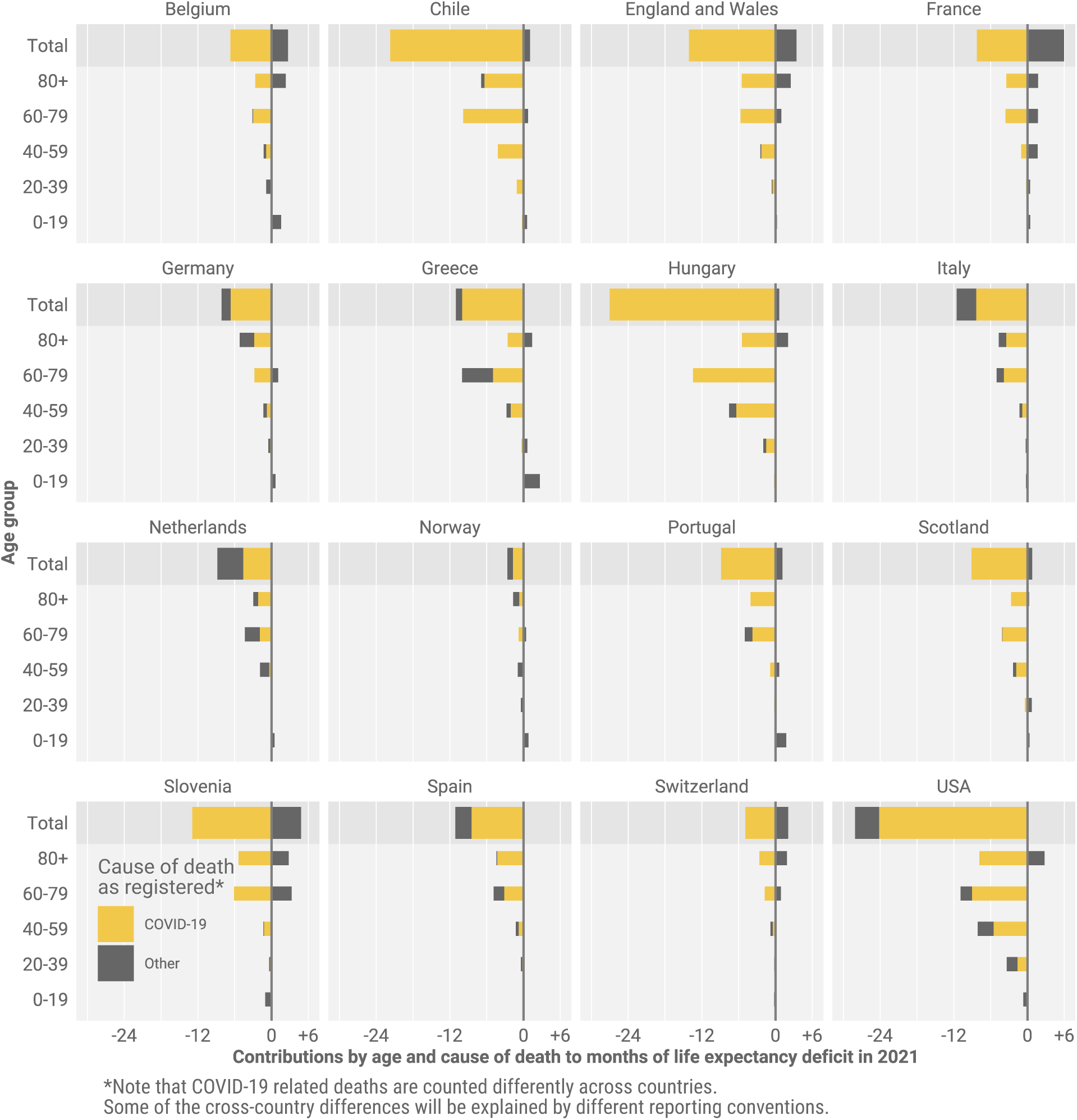
Female life expectancy deficit in 2021 decomposed into contributions by age and cause of death. LE deficit is defined as observed minus expected life expectancy had pre-pandemic mortality trends continued.

**Figure 14:**
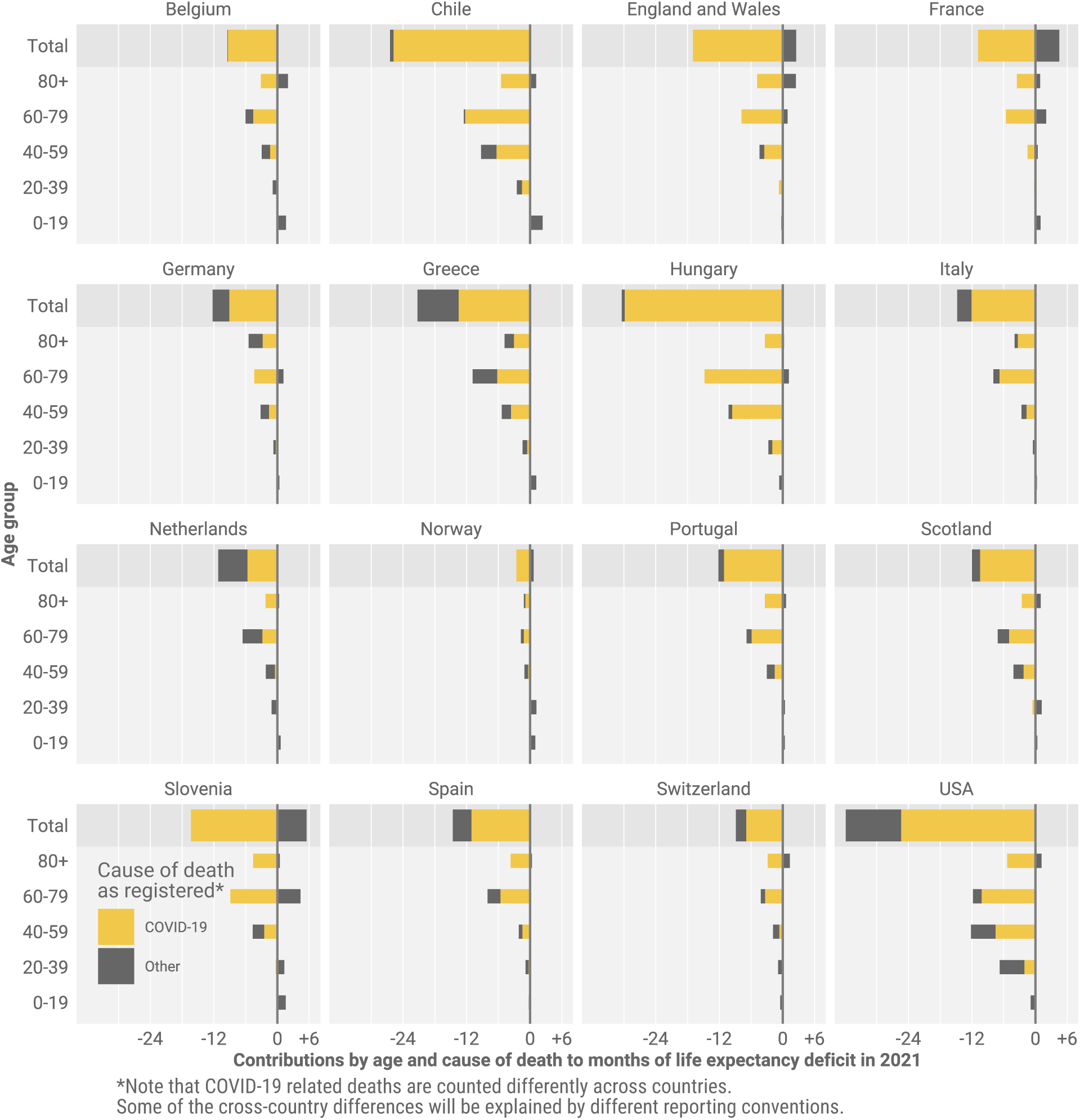
Male life expectancy deficit in 2021 decomposed into contributions by age and cause of death. LE deficit is defined as observed minus expected life expectancy had pre-pandemic mortality trends continued.

### Historic life expectancy losses

**Table 5:**
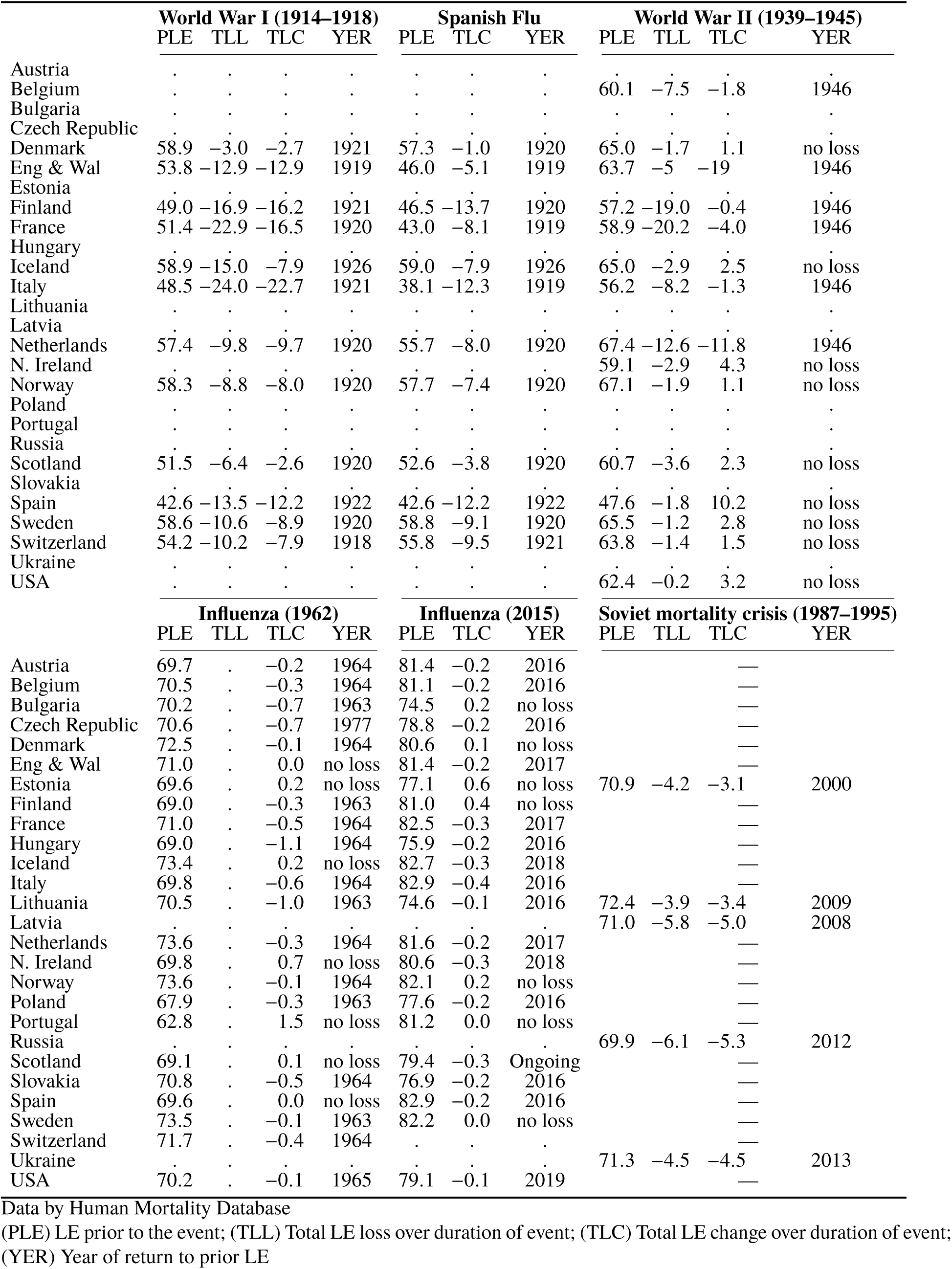
Life expectancy losses and bounce-backs during six selected mortality shock events in the 20th century.

